# Antidepressant Trial Duration versus Duration of Real-World Use: A Systematic Analysis

**DOI:** 10.1101/2025.02.27.25323057

**Authors:** William Ward, Alyson Haslam, Vinay Prasad

**Author notes:** **Corresponding author:** Vinay Prasad, Department of Epidemiology and Biostatistics, UCSF Mission Bay Campus | Mission Hall: Global Health & Clinical Sciences Building |. Disclosure: V.P. receives research funding from Arnold Ventures through a grant made to UCSF, and royalties for books and writing from Johns Hopkins Press, MedPage, and the Free Press. He declares consultancy roles with UnitedHealthcare and OptumRX; He hosts the podcasts, Plenary Session, VPZD, Sensible Medicine, writes the newsletters, Sensible Medicine, the Drug Development Letter and VP’s Observations and Thoughts, and runs the YouTube channel Vinay Prasad MD MPH, which collectively earn revenue on the platforms: Patreon, YouTube and Substack.

## Abstract

**Importance:** Antidepressant use is rising globally, with increasing duration of real-world prescribing. While the FDA considers 6-8 week trials adequate for regulatory approval, guidelines recommend prolonged treatment. This raises questions about the evidence supporting long-term prescribing practices.

**Objective:** To systematically compare the duration of placebo-controlled randomized trials of commonly prescribed antidepressants with real-world usage patterns.

Design, Setting, and Participants: This descriptive review analyzed 52 eligible placebo-controlled randomized trials (n=10,116 participants) investigating the 10 most commonly prescribed antidepressants, selected based on 2022 United States prescription data. Trials were sampled at 5-year intervals from 1978 through 2023.

Main Outcomes and Measures: The primary outcome was the comparison between trial duration and real-world antidepressant use duration based on National Health and Nutrition Examination Survey (NHANES) data. Secondary outcomes included methodological characteristics such as the use of standardized severity scales, withdrawal monitoring, taper protocols, and type of placebo used.

**Results:** The median duration of antidepressant use in the United States was approximately 5 years (260 weeks), while the median trial duration was 8 weeks (IQR: 6-12 weeks). Among trials, 88.5% (n=46) had a duration of 12 weeks or less, and only 11.5% (n=6) randomized participants beyond 12 weeks, with none exceeding 52 weeks. Although 94.2% of antidepressant users are prescribed medication for longer than 60 days, the median trial duration was 56 days. Few trials monitored for withdrawal symptoms (3.8%) or included taper protocols (18.9%), and only 1.9% reported depression or anxiety outcomes during the post-treatment period. No trials used active placebos to mitigate unblinding.

**Conclusions and Relevance:** A substantial discordance exists between the typical 8-week duration of clinical trials and the median 5-year real-world use of antidepressants. This gap, compounded by inadequate monitoring for withdrawal effects and post-treatment outcomes, raises important questions about the evidence supporting current long-term prescribing practices. Publicly funded trials of longer duration that monitor for withdrawal, sexual side effects, and relapse are necessary to determine optimal antidepressant therapy duration.

## Introduction

Antidepressant use is rising in the United States^1,2^ and across the globe.^3^ This trend reflects changes in both the duration and frequency of prescribing. Many trials evaluating the efficacy of antidepressants use a treatment duration of about 8-12 weeks.^4,5^ However, this duration is discordant with the duration of typical use.^6,7^ This difference raises questions about whether the available evidence sufficiently supports contemporary prescribing practices.

Antidepressants include serotonin reuptake inhibitors (SSRIs), serotonin-norepinephrine reuptake inhibitors (SNRIs), tricyclic antidepressants, and atypical antidepressants. Randomized controlled trials show that antidepressants are superior to placebo in the treatment of depressive^4^ and anxiety^8^ disorders. According to guidelines issued in 2018, the FDA considers antidepressant trials of 6-8 weeks to be adequate for regulatory approval.^9^

The duration of these trials is in contrast to treatment guidelines from the American Psychiatric Association (APA). In 2010, the organization recommended that, after acute treatment lasting 4 to 8 weeks, antidepressant therapy should continue for an additional 4 to 9 months. These guidelines also recommend indefinite treatment for certain populations as “maintenance therapy”.^10^

The APA justifies long-term therapy based on a discontinuation trial showing recurrent depression in 8% of those who continued long-term venlafaxine therapy versus 44% of participants who were randomized to placebo.^11^ However, it is unclear whether such trials adequately support long-term therapy.^12^ First, logically, whether withdrawing a drug is associated with harm is different than whether the initial long-term use was associated with sustained clinical benefit. Second, these trials fail to distinguish relapse from antidepressant withdrawal. Both syndromes lead to increased scores on the commonly used Montgomery–Åsberg Depression Rating Scale (MADRS) and Hamilton Rating Scale for Depression (HAMD).^13,14^ Given that discontinuation trials do not randomize participants at treatment onset, they cannot answer whether long-duration, short-duration, or no treatment results in the best long-term outcomes.

Trials that randomize participants at treatment onset are better suited to illustrate the benefits and harms of long-term antidepressant prescribing. A 2008 meta-analysis identified long-duration placebo-controlled randomized-controlled trials of various SSRIs. Six trials of six to eight month duration were identified. Antidepressant use of this duration was associated with increased response, but not remission. However, this finding was limited by study heterogeneity (I^2^=63.9%), high dropout rates (average 48%), and lack of withdrawal monitoring at trial completion.^15^

Since the publication of the 2008 meta-analysis, the need for robust long-term antidepressant data has become more apparent. According to the National Health and Nutrition Examination Survey (NHANES) of 2005-2008 and 2011-2014, the proportion of individuals taking antidepressants for 10 years or more in the United States increased from 13.6% to 25.3%.

Meanwhile, the proportion of individuals taking antidepressants in an acute setting (<60 days) decreased from 8.4% to 5.8% (See Figure 1).^6,7^

**Figure 1.**
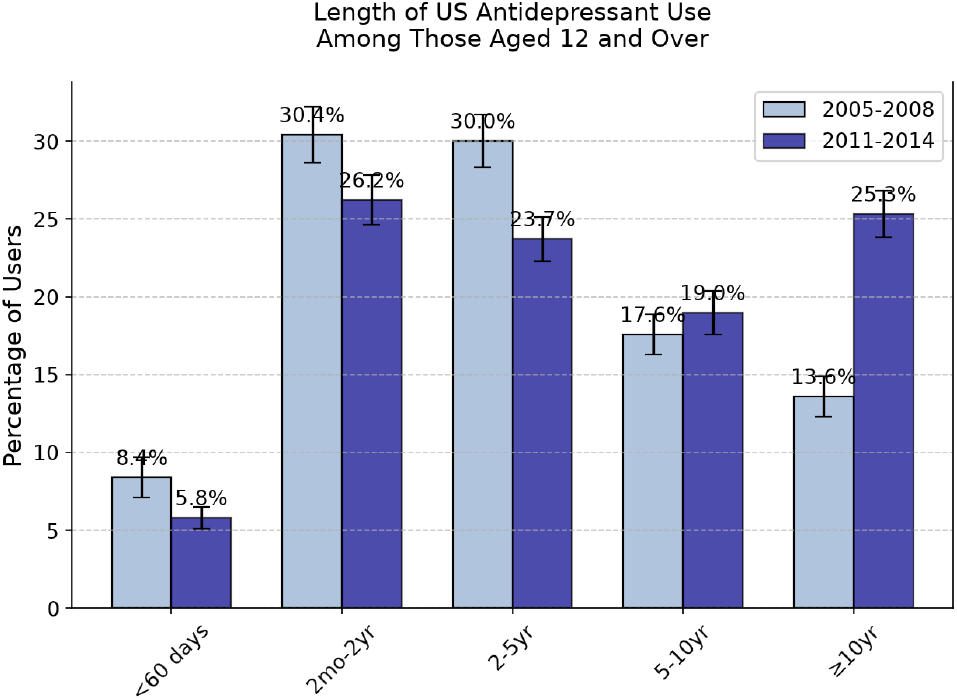
Duration of Antidepressant Use in the United States, 2005-2014. Data from the National Health and Nutrition Examination Survey (NHANES) show the percentage of antidepressant users aged ≥12 years by duration of use.^6,7^ Error bars indicate reported standard errors. A notable shift occurred between survey periods, with long-term use (≥10 years) increasing from 13.6% in 2005-2008 to 25.3% in 2011-2014, while short-term use (<60 days) decreased from 8.4% to 5.8%.

**Figure 2.**
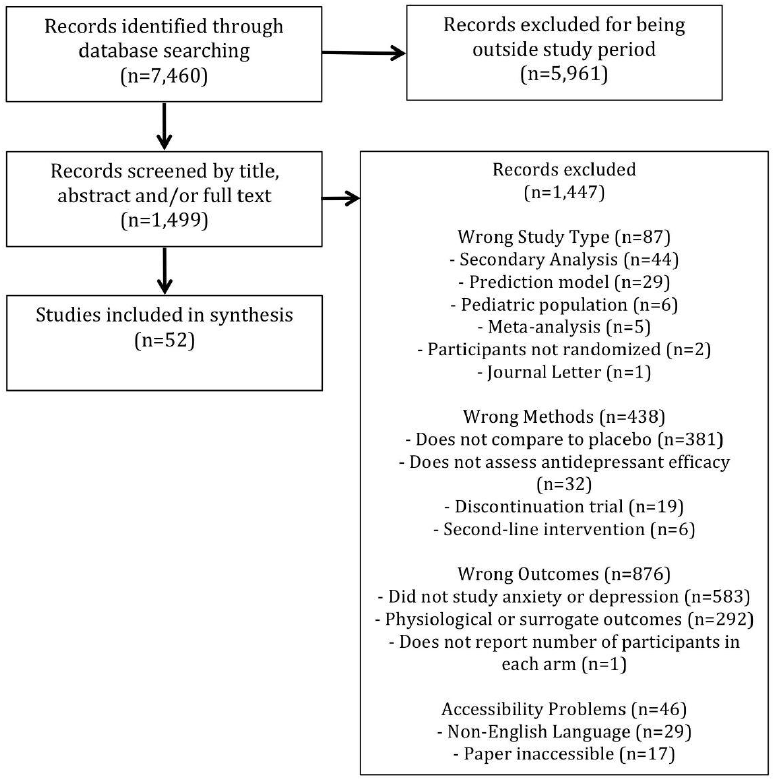
Duration Distribution of Randomized Clinical Trials for Antidepressants. The duration of 52 clinical trials is plotted in ascending order. Each bar represents a single trial, with trial duration shown on the vertical axis. The median duration was 8 weeks (red dashed line) and the mean duration was 11.0 weeks (blue dashed line). Only 3 trials (5.8%) exceeded 26 weeks.

**Figure 3.**
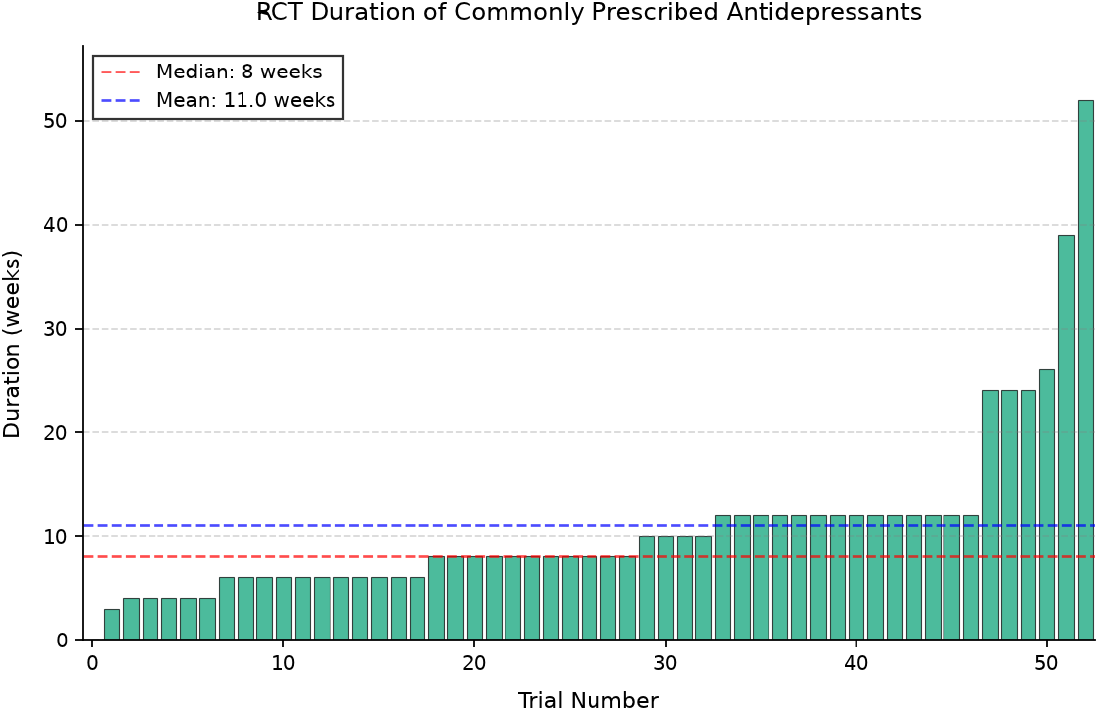

**Figure 4.**
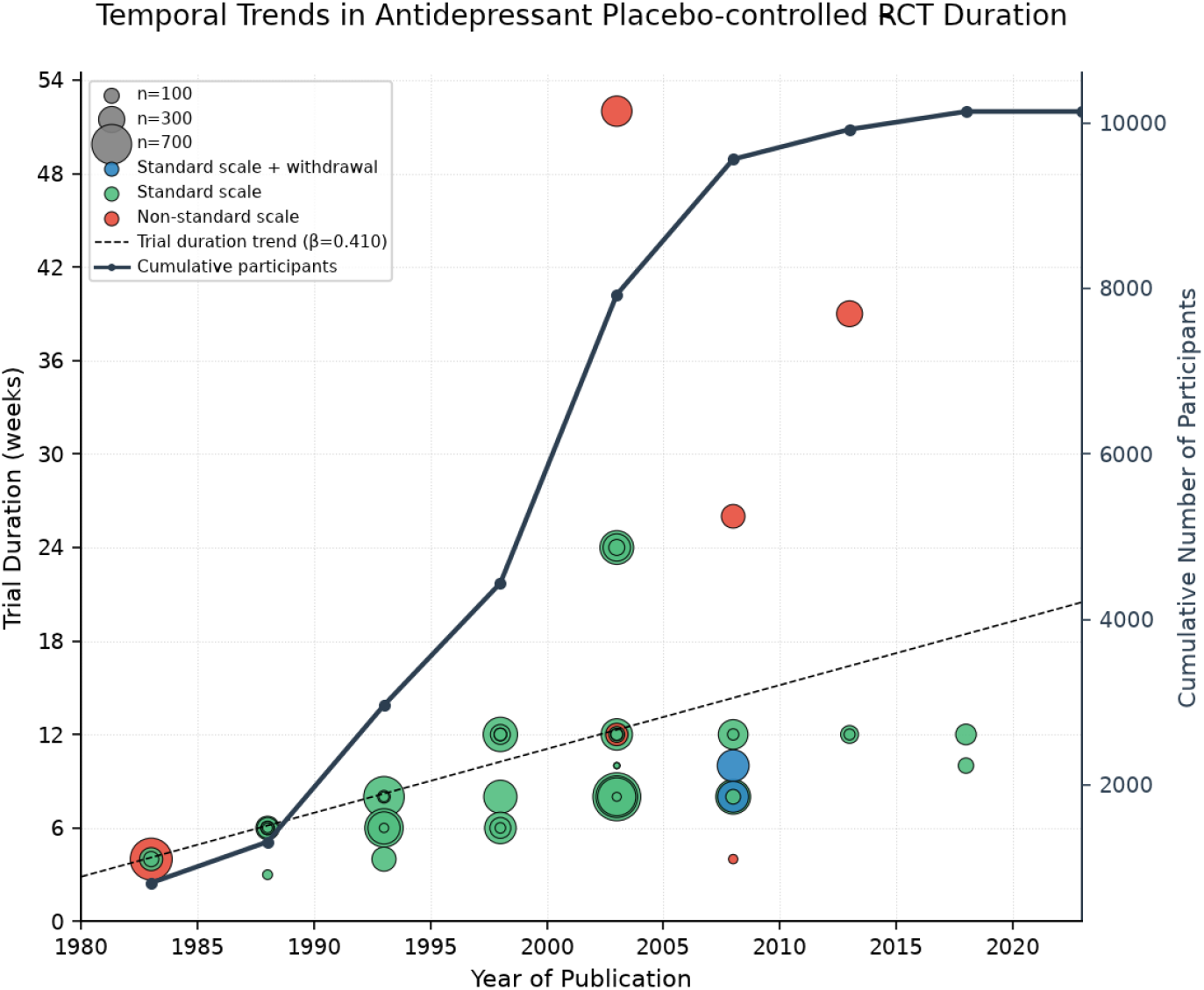
Temporal Trends in Antidepressant Placebo-controlled RCT Duration, 1983-2023. Circle size indicates sample size (range: 12-747 participants). Studies were classified by assessment method: standard severity scores such as HAMD, MADRS, or HAMA reported (green); standard depression rating scales with withdrawal monitoring (blue); or non-standard scales and no withdrawal monitoring (red). The dashed line shows the linear trend (β=0.41 weeks/year, R^2^=0.17, P=0.003). Mean trial duration was 11.0 weeks (median 8.0 weeks). Total participants across all studies: 10,116.

**Figure 5.**
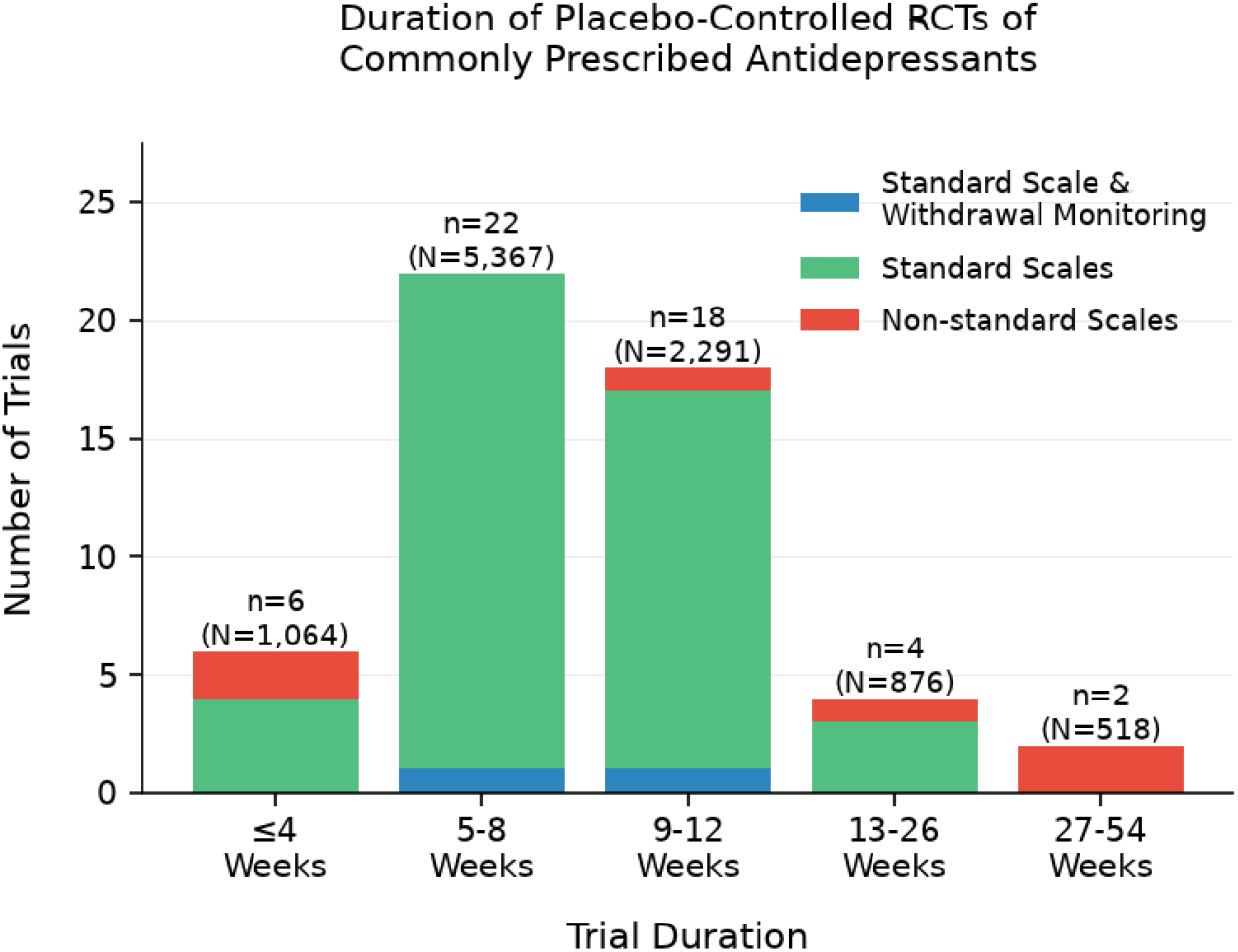
Duration of Antidepressant Clinical Trials. Distribution of 52 placebo-controlled randomized trials by duration (weeks), categorized by use of standard scales and withdrawal monitoring. Numbers above bars indicate total trials (n) and participants (N) in each duration category. Most trials (31/51) were 8 weeks or less in duration, with few extending beyond 26 weeks.

There is a growing need to identify the hazards and benefits of long-term prescribing. It is known that prolonged treatment duration and high antidepressant doses are associated with increased risk of withdrawal upon discontinuation,^16^ a risk documented as early as 1993.^17^ The APA claims these symptoms do not require treatment and resolve within two weeks.^10^ However, a systematic review found that withdrawal symptoms can be debilitating, extend beyond several months, and occur among 55% of those discontinuing antidepressants.^18^ Despite long-standing recognition of withdrawal, it is unclear whether monitoring for this syndrome is an established feature of antidepressant trials.

To better evaluate the discordance between the duration of antidepressant therapy in the real-world compared to clinical trials, we sought to systematically review the literature for clinical trials evaluating the efficacy of antidepressants and compare this with nationally reported usage patterns.

## Methods

This descriptive review aims to characterize the differences between antidepressant trial durations and real-world usage patterns. Our primary objective is to describe the duration of the placebo-controlled randomized trials that form the evidence base of the ten most commonly prescribed antidepressants. We compare these findings to the duration of antidepressant use according to NHANES. Other objectives include description of the temporal trends in trial duration and methodological characteristics such as the use of standard severity scales (HAMD, MADRS, HAMA); withdrawal monitoring; taper protocols; and type of placebo used.

We conducted a literature search of the 10 most commonly prescribed antidepressant medications (see Table 1), which includes SSRIs, SNRIs, tricyclic antidepressants, and atypical antidepressants. These medications were selected based on 2022 United States prescription data to ensure our analysis reflected contemporary prescribing patterns.^19^ We included trials that assess the efficacy of antidepressants in the treatment of either of the two conditions for which antidepressants are commonly prescribed: unipolar depression or generalized anxiety.

**Table 1.** Commonly prescribed antidepressants according to 2022 United States prescription data^19^.

| Rank Across All Drugs | Generic Name | Total Prescriptions | Users |
| --- | --- | --- | --- |
| 11 | Sertraline | 39,891,990 | 8,422,729 |
| 15 | Escitalopram | 30,750,612 | 7,493,104 |
| 18 | Trazodone | 27,363,318 | 5,759,352 |
| 21 | Bupropion | 25,197,923 | 6,111,736 |
| 22 | Fluoxetine | 24,014,263 | 5,174,534 |
| 31 | Duloxetine | 18,066,468 | 4,042,916 |
| 40 | Citalopram | 15,047,663 | 3,540,540 |
| 44 | Venlafaxine | 13,881,022 | 2,844,672 |
| 87 | Amitriptyline | 7,887,977 | 1,675,744 |
| 92 | Paroxetine | 7,127,748 | 1,470,173 |

**Table 2.** Summary of trial characteristics. An asterisk (*) denotes a statistically significant difference between long and short-duration trials. Given the limited number of long-duration trials, few statistically significant differences were identified.

|  | Trials ≤12 weeks<br>(n=46) | Trials >12 weeks<br>(n=6) | Total<br>(n=52) |
| --- | --- | --- | --- |
| Trial Characteristics |  |  |  |
| Total Participants | 8,722 | 1,394 | 10,116 |
| Mean trial duration (weeks) | 8.4 | 31.5 | 11.0 |
| Trial duration range (weeks) | 3-12 | 24-52 | 3-52 |
| Methodology Features |  |  |  |
| Standard severity scales used<br>(HAMD, MADRS, or HAMA) | 43* | 3* | 46 |
| Active Placebo Used | 0 | 0 | 0 |
| Taper Protocol Included | 7 | 2 | 9 |
| Withdrawal Monitoring | 2 | 0 | 2 |
| Relapse monitoring | 2 | 0 | 2 |
| Post-trial open-label treatment | 3 | 1 | 4 |
| Population Characteristics |  |  |  |
| Weighted Average Age (years) | 49.0 | 55.0 | 49.9 |
| Range of ages (years) | 16-97 | 19-98 | 16-98 |
| Female participants (%) | 58.8% | 63.6% | 59.4% |
| Trials requiring co-morbid condition | 16 | 4 | 20 |

### Search strategy

We searched PubMed for “Duloxetine OR Cymbalta OR Citalopram OR Celexa OR Sertraline OR Zoloft OR Fluoxetine OR Prozac OR Trazodone Or Desyrel OR Escitalopram OR Lexapro OR Paroxetine OR Paxil OR “Paxil CR” OR Venlafaxine OR Effexor OR Effexor XR OR bupropion OR Wellbutrin” clinical trials or observational studies involving adult (19+) human subjects. We extracted publications at 5-year intervals to obtain a representative temporal sample from 1978 through 2023 (1978, 1983, 1988, 1993, 1998, 2003, 2008, 2013, 2018, and 2023). Relevant studies were identified by two authors (W.W. and A.H.). The most recent search was made on 12/5/2024. For a full list of searches, please see supplemental materials.

### Inclusion and exclusion criteria

Studies were included if participants were diagnosed with generalized anxiety and/or unipolar depression (i.e. dysthymia, major depressive disorder, or major depressive episode). Studies were included if participants were randomized to placebo or antidepressants as first-line monotherapy and if the study followed endpoints for disease progression, such as severity, response, or remission. Studies were included regardless of diagnostic criteria, severity scoring system, or setting. We did not exclude studies based on comorbid conditions, such as coronary artery disease or dementia. Studies were excluded for the following reasons: the design was a meta-analysis, secondary analysis, or review article; participants were predominantly younger than 18 years of age; endpoints were limited to biomarkers, drug levels, or imaging changes; the primary condition evaluated was not an anxiety or depressive disorder; participants were randomized to discontinue an antidepressant after non-randomized use; the control group was provided a treatment intervention (e.g. medication, psychotherapy, exercise, etc) not provided to the medication arm.

### Data Extraction And Analysis

From each study, we extracted the medication studied, duration of trial, which outcomes were measured, and placebo specifications. For trials that randomized participants to three or more arms, data were not extracted from alternative treatment interventions. We also extracted inclusion criteria; total number of participants; number in each study arm; and demographic information such as average age, range of ages, comorbid conditions, and percentage of female participants. We also collected details regarding adverse event reporting, taper protocols, and withdrawal monitoring. We also determined whether a trial’s protocol included follow-up upon trial completion for relapse monitoring.

We systematically evaluated features that make trials useful for meta-analytic purposes and to guide real-world prescribing patterns. For each trial, we identified which severity measures were used (e.g. HAMD, MADRS, or HAMA).

To compare the duration of antidepressant trials to typical prescribing, the most recently published NHANES survey data were used. NHANES collects medication usage data through household interviews where participants are asked to report all prescription medications taken in the past 30 days. Duration of use was determined through participant self-report. Duration categories include <60 days, 60 days-2 years, 2-5 years, 5-10 years, and ≥10 years, as previously reported.^6,7^

### Statistical analysis

Our results are largely descriptive and presented in frequencies (percentages) and medians (ranges). All analyses were done in R version 4.2.0 (R Foundation for Statistical Computing, Vienna, Austria).

Because our study involved publicly available data and did not involve individual patient data, this study was not submitted to the institutional review board.

## Results

We identified 52 eligible placebo-controlled randomized trials (n=10,827) investigating the ten most commonly prescribed antidepressants. After excluding trial arms with interventions not included in our analysis (e.g., alternative treatments), the final analysis included 10,116 participants. The median duration of antidepressant use in the United States was approximately 5 years (260 weeks), while the median trial duration was 8 weeks. While 94.2% of those taking antidepressants have received prescriptions for longer than 60 days, the median antidepressant placebo-controlled randomized-controlled trial is 56 days.

Among all trials, two trials (3.8%) monitored for withdrawal symptoms, nine (17.3%) included taper protocols, and two (3.8%) reported depression or anxiety relapse during the post-treatment period. 13 trials (25%) specifically monitored sexual side effects (e.g. decreased libido, erectile dysfunction, etc).

Trial duration ranged from 3 to 52 weeks, with a median of 8 weeks (IQR: 6-12 weeks). 88.5% of trials (n=46) had a duration of 12 weeks or less. 94.1% of trials with a duration shorter than 8 weeks (n=17) were published prior to 2000, while 83.3% of trials with a duration longer than 8 weeks (n=24) were published after 2000. This association between trial duration and publication period was statistically significant (χ^2^ = 23.9, p < 0.001). Six trials (11.5%) randomized participants beyond 12 weeks, with durations of 24 (n=3), 26, 39, and 52 weeks. No trials randomized participants to different treatment durations.

Among trials of 3-12 week duration, 42 of 45 (93.2%) used at least one standard depression or anxiety severity scale such as HAMD, MADRS, or HAMA. Of the six trials exceeding 12 weeks, only three used standard scales. Of the trials that comprehensively reported both response and remission outcomes, ten were trials of 8-12 week duration and one was a trial of 24 week duration.

Monitoring for antidepressant withdrawal was uncommon. Two trials—of 10 and 12-week duration—monitored for “discontinuation-emergent adverse events”.^20,21^ Neither trial utilized a specific time horizon or survey for monitoring. More often, trials offered open-label maintenance therapy upon trial completion, a feature that appears in five trials. No trials utilized an active placebo to mitigate unblinding.

The included trials investigated the effect of antidepressants on depression (n=48, 90.6%), anxiety (n=4, 7.5%), and mixed anxiety/depression (n=1, 2.1%). 20 trials (37.7%) recruited participants with a specific comorbid condition.

## Discussion

The analysis revealed a striking disparity between the duration of clinical trials and real-world use patterns. According to NHANES data,^7^ 25% of antidepressant users have taken them for over 10 years. Given 13.2% of U.S. adults take antidepressants^22^ and current census figures^23^, this represents approximately 8.8 million adults. Meanwhile, few clinical trials testing efficacy evaluate outcomes beyond 12 weeks of therapy. While the FDA considers short-duration trials sufficient for regulatory approval, antidepressants are increasingly used for indefinite periods of time. Withdrawal and relapse monitoring at treatment completion would help validate the optimal duration of treatment in clinical trials. However, these features are missing from clinical trials of all durations, even though the risk of withdrawal has been known for 30 years. Our analysis found that only 3.8% of trials monitored for withdrawal symptoms, 18.9% included taper protocols, and just 1.9% reported post-treatment outcomes. These methodological limitations severely restrict our ability to make informed decisions about long-term prescribing.

Those that experience withdrawal upon antidepressant discontinuation experience symptoms of variable duration and severity.^24^ Because of this heterogeneity of experiences, placebo-controlled randomized trials present a valuable opportunity to better understand risk factors for withdrawal and other hazards of long-term antidepressant use. Given that such data should weigh on decisions to initiate or continue antidepressant therapy, withdrawal monitoring should be included and standardized in future antidepressant trials using a validated severity scale such as the Discontinuation-Emergent Signs and Symptoms checklist.^25^

Several long-term trials were identified in this study, yet methodological shortcomings limit their generalizability. The few trials that extended beyond 12 weeks utilized strict inclusion criteria and atypical measures of disease severity. For example, four of six long-term trials required a specific co-morbid condition such as acute coronary syndrome, breast cancer, alcohol use disorder in remission, or Alzheimer’s disease. Three of the long-duration trials failed to use a standard measure of anxiety or depression such as MADRS, HAMD or HAMA, making published data unhelpful for meta-analytic purposes.

Considering NHANES^7,22^ and census data^23^, the number of adults who have taken antidepressants for longer than 2 years represents approximately 23.9 million people. Yet, our cross-sectional study did not capture any trials longer than one year.

In the absence of long-term data, prescribers must rely upon short-term trials. While there is a slight trend of trial prolongation, this is on the scale of weeks over the course of multiple decades. There is a critical need for long-term efficacy and safety data from randomized trials. Until long-term trials are done, the duration of antidepressant prescribing is based upon indirect measures of efficacy and safety such as discontinuation trials, now known to be flawed. This disconnect has real-world consequences. Prolonged use is associated with increased misdiagnosis, healthcare utilization, long-term dependency, and severity of withdrawal symptoms.^26^ As is the case with benzodiazepines, indefinite antidepressant use may produce diminishing benefits and increasing hazards.

Results from RCTs are strengthened when every opportunity is taken to mitigate unblinding. One meta-analysis suggests that the comparison of an inert placebo and antidepressant can result in unblinding. An active placebo is a non-therapeutic substance with a side-effect profile similar to the treatment arm used to prevent unblinding. The benefit of tricyclic antidepressants was found to be less when compared to active placebos compared to an inert placebo.^27^ While active placebos were occasionally used in tricyclic antidepressant trials in the 1960s and 1970s, no trials using an active placebo were identified in this study. To better quantify the psychotropic benefit of antidepressants, active placebos should be considered in future antidepressant trials of all durations.

This systematic review has several important limitations. First, the strategy of extracting publications at 5-year intervals may have resulted in important trials being overlooked in intervening years. A dataset that includes all years would allow for better analysis of temporal trends and differences between long and short-duration trials.

Secondly, this review only included placebo-controlled trials, which may have shorter durations of therapy than active-comparator trials. However, we used this selection criterion because it better selected trials that established absolute efficacy. Third, our comparison of real-world usage relies upon NHANES data from 2011-2014, which is now a decade old. Given trends in antidepressant prescribing continue to evolve, more recent prescribing data would be valuable to update comparisons and guide antidepressant trials. Despite these limitations, our findings demonstrate a significant deficiency of long-duration trials.

## Conclusion

The evidence base of antidepressants, particularly their duration of use, is an important public health question as 13.2% of adults are taking antidepressants.^7^ The efficacy of commonly prescribed antidepressants is largely based on trials of 8-12 week duration, and yet, we find, the median duration of therapy in the real world is 5 years. In this cross-sectional study, few placebo-controlled RCTs extended beyond six months and none beyond 1 year. Trials with a duration longer than 12 weeks suffer from limitations (e.g., specific study populations) that prevent generalizability.

Regardless of duration, trials rarely monitored participants for withdrawal or relapse upon completion of treatment. Additionally, no trials attempted to prevent unblinding with the use of an active placebo. Publicly funded randomized controlled trials comparing antidepressants to placebo with long duration, which also monitor for withdrawal, sexual side effects and relapse upon treatment discontinuation are necessary to determine the optimal duration of therapy.

## Data Availability

All data produced in the present study are available upon reasonable request to the authors

## Notes

### Funding Statement

This study did not receive any funding

### Author Declarations

- Government publication (e.g., NHANES data from the CDC) - Previously published literature (the 52 placebo-controlled randomized trials you analyzed)

