## Supplemental Table 1 & 2 for "Antidepressant Trial Duration versus Duration of Real-World Use: A Systematic Analysis"

#### Supplementary Appendix

### Contents

**Supplementary Table 1: Characteristics of Included Trials** **3**
**Supplementary Table 2:  Outcome Measures and Monitoring** **8**

### Supplementary Table 1: Characteristics of Included Trials

| Study Name | Trial Duration (weeks) | No. of arms included | Total study arms | Drug | No. randomized | Inclusion Diagnosis | Mean Age (y) | Age Range (y) | % Female | Comorbidity |
|---|---|---|---|---|---|---|---|---|---|---|
| Claghorn (1983) | 4 | 2 | 3 | amitriptyline | 85 | MDD (RDC) | 39 | 18-65 | 57% | |
| | | | | placebo | 87 | | | | | |
| Zung (1983) | 4 | 2 | 2 | bupropion | 48 | MDD (DSM II) | 41 | 18-70 | 23% | Hospitalized |
| | | | | placebo | 27 | | | | | |
| Downing (1983) | 4 | 2 | 2 | amitriptyline | 281 | depression, unspecified | 44 | 18-70 | 76% | |
| | | | | placebo | 288 | | | | | |
| Paykel (1988) | 6 | 2 | 2 | amitriptyline | 67 | Major & Minor Depression (RDC) | 38 | 18-64 | 83% | |
| | | | | placebo | 74 | | | | | |
| Klieser (1988) | 3 | 3 | 3 | amitriptyline | 11 | MDD (DSM III) | 42 | | 69% | |
| | | | | trazodone | 10 | | | | | |
| | | | | placebo | 10 | | | | | |
| Hicks (1988) | 6 | 2 | 2 | amitriptyline | 16 | Major depression with melancholia (DSM-III) | 42 | 18-59 | | |
| | | | | placebo | 15 | | | | | |
| Byerley (1988) | 6 | 2 | 2 | fluoxetine | 32 | Major depression (DSM-III) | 39 | | 68% | |
| | | | | placebo | 29 | | | | | |
| Hollyman (1988) | 6 | 2 | 2 | amitriptyline | 90 | Depression (RDC) | | 18-64 | 83% | |
| | | | | placebo | 88 | | | | | |
| Muijen (1988) | 6 | 2 | 2 | fluoxetine | 26 | Major Depressive Illness (RDC) | 35 | | 63% | |
| | | | | placebo | 28 | | | | | |
| Heiligenstein (1993) | 8 | 2 | 2 | fluoxetine | 24 | Major depression (DSM-III-R) | 42 | 18-65 | 65% | |
| | | | | placebo | 28 | | | | | |
| Tollefson (1993) | 8 | 2 | 2 | fluoxetine | 264 | Major depression (DSM-III-R) | 68 | 60+ | 55% | |
| | | | | placebo | 270 | | | | | |
| Katz (1993) | 4 | 2 | 2 | amitriptyline | 95 | Major depression (DSM-III) | 44 | 18-70 | 43% | |

| | | | | | | | | | | |
|---|---|---|---|---|---|---|---|---|---|---|
| | | | | placebo | 94 | | | | | |
| Dunbar (1993) | 6 | 2 | 2 | paroxetine | 167 | MDD (DSM-III) | 41 | 18-65 | 51% | |
| | | | | placebo | 169 | | | | | |
| Feighner (1993) | 6 | 2 | 3 | paroxetine | 240 | MDD (DSM-III) | 40 | 18-65 | 53% | |
| | | | | placebo | 240 | | | | | |
| Hellerstein (1993) | 8 | 2 | 2 | fluoxetine | 16 | Dysthymia (DSM-III-R) | 36 | 21-65 | 50% | |
| | | | | placebo | 16 | | | | | |
| Kusalic (1993) | 6 | 2 | 2 | amitriptyline | 13 | MDE (DSM-III) | 41 | 22-61 | 37% | |
| | | | | placebo | 15 | | | | | |
| Khan (1998) | 12 | 2 | 2 | venlafaxine 75mg/day | 96 | MDE (DSM-III) | 42 | 18+ | 64% | |
| | | | | venlafaxine 150mg/day | 96 | | | | | |
| | | | | venlafaxine 200mg/day | 94 | | | | | |
| | | | | placebo | 98 | | | | | |
| Elliott (1998) | 12 | 2 | 2 | paroxetine | 25 | MDE (DSM-III) | 36 | | 7% | HIV positive |
| | | | | placebo | 25 | | | | | |
| Rudolph (1998) | 6 | 2 | 2 | venlafaxine 75mg/day | 77 | MDE (DSM-III) | 43 | 18-65 | 36% | |
| | | | | venlafaxine 225 mg/day | 79 | | | | | |
| | | | | venlafaxine 375 mg/day | 75 | | | | | |
| | | | | placebo | 92 | | | | | |
| Petrakis (1998) | 12 | 2 | 2 | fluoxetine | 23 | MDE (DSM-III-R) | 34 | | 36% | Opioid dependent |
| | | | | placebo | 31 | | | | | |
| Reimherr (1998) | 8 | 2 | 2 | bupropion SR 150mg daily | 121 | MDE (DSM-III) | 39 | 18-72 | 70% | |
| | | | | bupropion SR 150mg BID | 120 | | | | | |
| | | | | placebo | 117 | | | | | |
| Kramer (1998) | 6 | 2 | 2 | paroxetine | 72 | MDD (DSM-IV) | | 18-65 | | |
| | | | | placebo | 70 | | | | | |
| Roy (1998) | 6 | 2 | 2 | sertraline | 18 | MDE (DSM-III) | 41 | | 8% | Recently abstinent alcoholics |
| | | | | placebo | 18 | | | | | |

| | | | | | | | | | | |
|---|---|---|---|---|---|---|---|---|---|---|
| Fava (1998) | 12 | 3 | 3 | paroxetine | 55 | Depression (Raskin Depression Scale score ≥8 and a HAMD score ≥18) | 41 | | 51% | |
| | | | | fluoxetine | 54 | | | | | |
| | | | | placebo | 19 | | | | | |
| Patel (2003) | 52 | 2 | 2 | fluoxetine | 150 | Mixed anxiety depressive disorder (43%) and depressive disorder (53%) (ICD-10) | 49 | 48.6 | 83% | |
| | | | | placebo | 150 | | | | | |
| Fruehwald (2003) | 12 | 2 | 2 | fluoxetine | 26 | Post-stroke depression (Hamilton Depression Score > 15) | 64 | 25-85 | 42% | Stroke |
| | | | | placebo | 24 | | | | | |
| Rickels (2003) | 8 | 3 | 3 | paroxetine 20mg | 189 | GAD (DSM-IV) | 40 | 18+ | 55% | |
| | | | | paroxetine 40 mg | 197 | | | | | |
| | | | | placebo | 180 | | | | | |
| Mulholland (2003) | 8 | 2 | 2 | sertraline | 13 | Depressive symptoms (BDI ≥15, BPRS depression item ≥3) | 38 | | 38% | Chronic schizophrenia |
| | | | | placebo | 13 | | | | | |
| Fisch (2003) | 12 | 2 | 2 | fluoxetine | 83 | Depression (BZSDS) | 60 | | 50% | Advanced cancer (various types) |
| | | | | placebo | 80 | | | | | |
| Paile-Hyvärinen (2003) | 10 | 2 | 2 | paroxetine | 7 | Mild depression (MADRS score 2.5-12) | 62 | | 100% | Post-menopausal, T2DM |
| | | | | placebo | 6 | | | | | |
| Leentjens (2003) | 10 | 2 | 2 | sertraline | 6 | MDD (DSM-IV) | 67 | | 33% | Parkinson's Disease |
| | | | | placebo | 6 | | | | | |
| Lepola (2003) | 8 | 3 | 3 | citalopram | 160 | MDD (DSM-IV) | 44 | | 72% | |
| | | | | escitalopram | 155 | | | | | |
| | | | | placebo | 154 | | | | | |
| Schneider (2003) | 8 | 2 | 2 | sertraline | 371 | MDD (DSM-IV) | 70 | 59-97 | 56% | |
| | | | | placebo | 376 | | | | | |

| | | | | | | | | | | |
|---|---|---|---|---|---|---|---|---|---|---|
| Lyketsos (2003) | 12 | 2 | 2 | sertraline | 24 | MDD (DSM-IV) | 78 | | 68% | Probable Alzheimer's disease |
| | | | | placebo | 20 | | | | | |
| Lenox-Smith (2003) | 24 | 2 | 2 | venlafaxine XL | 122 | GAD (DSM-IV) | 47 | 19-79 | 59% | |
| | | | | placebo | 122 | | | | | |
| Moak (2003) | 12 | 2 | 2 | sertraline | 38 | MDD or dysthymia (DSM-III-R) | 42 | | 39% | Alcohol use disorder |
| | | | | placebo | 44 | | | | | |
| Gual (2003) | 24 | 2 | 2 | sertraline | 44 | MDD or dysthymia (DSM-IV) | 47 | | 47% | Alcohol use disorder |
| | | | | placebo | 39 | | | | | |
| Rapaport (2003) | 12 | 2 | 2 | paroxetine CR | 104 | MDD (DSM-IV) | 70 | 60-88 | 56% | |
| | | | | paroxetine IR | 106 | | | | | |
| | | | | placebo | 109 | | | | | |
| Swenson (2003) | 24 | 2 | 2 | sertraline | 184 | MDD (DSM-IV) | 53 | | 37% | Acute coronary syndrome |
| | | | | placebo | 185 | | | | | |
| Rynn (2008) | 10 | 2 | 2 | duloxetine | 168 | GAD (DSM-IV) | 42 | 18+ | 62% | |
| | | | | placebo | 159 | | | | | |
| Bose/Korotzer (2008) | 8 | 3 | 3 | escitalopram | 127 | GAD (DSM-IV) | 38 | 18-65 | 62% | |
| | | | | venlafaxine | 129 | | | | | |
| | | | | placebo | 136 | | | | | |
| Navari (2008) | 26 | 2 | 2 | fluoxetine | 90 | "depressive symptoms" (TQSS score ≥ 2), excluded if "clinical depression" | 56 | 37-85 | 100% | Early-stage breast cancer prior to adjuvant therapy |
| | | | | placebo | 90 | | | | | |
| Kraus (2008) | 4 | 2 | 2 | citalopram | 14 | Interferon-associated depression, HADS >9 | 40 | 24-57 | 39% | Chronic hepatitis C |
| | | | | placebo | 14 | | | | | |
| Bose/Li (2008) | 12 | 2 | 2 | escitalopram | 130 | MDD (DSM-IV) | 68 | 60+ | 60% | |
| | | | | placebo | 134 | | | | | |
| Ehde (2008) | 12 | 2 | 2 | paroxetine | 21 | MDD and/or dysthymia (DSM-IV) | 45 | 24-63 | 52% | Multiple sclerosis |
| | | | | placebo | 21 | | | | | |

| | | | | | | | | | | |
|---|---|---|---|---|---|---|---|---|---|---|
| Raskin (2008) | 8 | 2 | 2 | duloxetine | 207 | MDD (DSM-IV) | 72 | 65-90 | 60% | |
| | | | | placebo | 104 | | | | | |
| Yonkers (2008) | 8 | 2 | 2 | paroxetine | 35 | MDD (DSM-IV) | 26 | 16+ | 100% | Postpartum state |
| | | | | placebo | 35 | | | | | |
| Levin (2013) | 12 | 2 | 2 | venlafaxine extended release | 51 | MDD or dysthymia (DSM-IV) | 35 | | 26% | Cannabis use |
| | | | | placebo | 52 | | | | | |
| Banerjee (2013) | 39 | 3 | 3 | sertraline | 107 | Depression in Dementia (Cornell Scale) & MDD (DSM-IV) | 79 | 47-98 | "majority" | Alzheimer's disease (NINCDS-ADRDA criteria) |
| | | | | placebo | 111 | | | | | |
| Ravindran (2013) | 12 | 2 | 2 | paroxetine | 21 | Dysthymic disorder (DSM-IV-TR) | 42 | 19-59 | 48% | |
| | | | | placebo | 19 | | | | | |
| Brown (2018) | 12 | 2 | 2 | escitalopram | 69 | MDD (DSM-IV) | 46 | 18-70 | 74% | Asthma |
| | | | | placebo | 70 | | | | | |
| Valle-Cabrera (2018) | 10 | 2 | 2 | sertraline | 39 | MDD (DSM-IV) | 45 | 19-62 | 92% | |
| | | | | placebo | 38 | | | | | |

**Supplementary Table 1 Abbreviations:** MDD: Major Depressive Disorder; MDE: Major Depressive Episode; RDC: Research Diagnostic Criteria; DSM-II/III/III-R/IV/IV-TR: Diagnostic and Statistical Manual of Mental Disorders (editions II, III, III-Revised, IV, IV-Text Revision); ICD-10: International Classification of Diseases, 10th Edition; GAD: Generalized Anxiety Disorder; SR: Sustained Release; CR: Controlled Release; IR: Immediate Release; XL/XR: Extended Release; BID: Twice daily; T2DM: Type 2 Diabetes Mellitus; HIV: Human Immunodeficiency Virus; NINCDS-ADRDA: National Institute of Neurological and Communicative Disorders and Stroke-Alzheimer's Disease and Related Disorders Association

### Supplementary Table 2: Outcome Measures and Monitoring

| Study Name | Surveys used | Primary Endpoint(s) | Reponse & Remission Outcomes Reported | Relapse Monitoring Upon RCT Completion | AEs Reporting w/ frequency and stat. analysis | Dropouts Reported | Sexual Dysfx Reporting | Taper Protocol | Withdrawal Monitoring | Post-trial open label treatment reported |
|---|---|---|---|---|---|---|---|---|---|---|
| Claghorn (1983) | HAMD 21, CGI-S, CGI-I | Change in Hamilton Depression Scale score | Response reported | No | Yes | Yes | No | No | No | Optional 6 month amitriptyline continuation |
| Zung (1983) | CGI-S, CGI-I, HAMD unspecified, HAMA, Zung Depression, Zung Anxiety | CGI-I, CGI-G, and Hamilton Rating Scale improvement | Response reported | No | Yes | Yes | No | No | No | No |
| Downing (1983) | Physician psychopathology rating scale 1-7 | Physician psychopathology rating scale 1-7 | Response reported | No | Not reported | No | No | No | No | No |
| Paykel (1988) | HAMD 17, Clinical Interview for Depression, Raskin Scale, CGI-I, CGI-S, Zung Depression, Zung Anxiety | Change in HAMD, CGI-S, CGI-I | Response reported | No | Limited AE reporting | Yes | No | No | No | No, but GP advised of treatment |
| Klieser (1988) | CGI, BPRS, HAMD unspecified, HAMA and AMDP | Change in HAM-D score | No | No | AEs reported elsewhere | Yes | No | No | No | No |

| | | | | | | | | | | |
|---|---|---|---|---|---|---|---|---|---|---|
| Hicks (1988) | HAMD unspecified, Carroll Rating Scale, Core Symptom Checklist, RADS, CAS, 7-point physician's global assessment scale, 7-point patient's global assessment scale, self-rating symptom scales | Change in HRSD score, global impression ratings | No | No | Yes | Yes | No | 2 week taper | No | No |
| Byerley (1988) | HAMD 21, CGI, global ratings of improvement | HAMD, CGI | No | No | Yes | Yes | No | No | No | No |

| | | | | | | | | | | |
|---|---|---|---|---|---|---|---|---|---|---|
| Hollyman (1988) | HAMD 17, CGI-S/I, Clinical Interview for Depression, Present State Examination, Newcastle Diagnostic Index, SCL, RADS | Change in Hamilton Depression Scale score, Clinical Interview for Depression, Raskin Three Area Depression Scale, Global ratings of severity and change | No | No | No | Yes | No | No | No | GP advised of treatment |
| Muijen (1988) | HAMD 17, MADRS, CGI | HDRS score improveme nt | Response reported | No | Yes | Yes | Yes, reported as “sexual dysfunction” | No | No | No |
| Heiligenstein (1993) | MADRS | MADRS response rate (≥50% reduction in score) | Response reported | No | Frequency without statistical analysis | Yes | Yes | No | No | No |
| Tollefson (1993) | HAMD 21 | HAMD score change | Yes | No | Yes | No | No | No | No | No |
| Katz (1993) | HAMD unspecified | HDRS total score change | No | No | No | No | No | No | No | No |

| | | | | | | | | | | |
|---|---|---|---|---|---|---|---|---|---|---|
| Dunbar (1993) | HAMD 17, MADRS, CGI, PGE | HAMD total score | Response reported | No | Yes | Yes | Freq. of decreased libido and abnormal ejaculation reported | No | No | No |
| Feighner (1993) | HAMD 17, MADRS, CGI, CAS, PGE | HAMD total score | Response reported | No | Yes | Yes | “Decreased libido” reported w/o detailed analysis | No | No | No |
| Hellerstein (1993) | HAMD 24, CGI, Cornell Dysthymia Rating Scale, SCL-58 | HAMD response and CGI score of "much improved" | Response reported | No | Frequency without statistical analysis | Yes | Frequency of “sexual dysfunction” reported | | No | |
| Kusalic (1993) | HAMD 17 | thyroid assay | No | No | No detailed AE reporting | No | No | No | No | No |
| Khan (1998) | HAMD 21, MADRS, CGI, HAM-D Anxiety-Psychic Item, HAM-D Anxiety-Somatization Factor | HAMD, MADRS total, CGI Scale | Response reported | No | Yes | Yes | No | No | No | No |
| Elliott (1998) | HAMD 21, HAMA, CGI, Brief | HAMD and CGI Scores, | Response reported | No | Yes | Yes | Yes | No | No | No |

| | | | | | | | | | | |
|---|---|---|---|---|---|---|---|---|---|---|
| | Symptom Inventory | HAMD response | | | | | | | | |
| Rudolph (1998) | HAMD 21, MADRS, CGI | HAM-D21, MADRS, and CGI scores | Response reported | No | Yes | Yes | No | Yes | No | No |
| Petrakis (1998) | HAMD 17, BDI, ASI, SCID | HDRS, BDI score change | No | No | Yes, limited | Yes | No | No | No | No |
| Reimherr (1998) | HAMD 17, CGI-S/I | HAM-D-17, CGI-S, and CGI-I scores | Response reported | No | Frequency of common AEs without statistical analysis | Yes | Yes | No | No | No |
| Kramer (1998) | HAMD 21, HAM-A, CGI-S | HAM-D21 score | Response reported | No | Yes | Yes | Yes | No | No | No |
| Roy (1998) | HAMD 21, BDI, CGI | HDRS Score improveme nt | Response reported | No | No | Yes | No | No | No | No |
| Fava (1998) | HAMD 21, RADS, CAS, Activity/Prod uctivity Assessment | HAMD-21, CAS | Response reported | No | Yes | Yes | Yes | No | No | No |

| | | | | | | | | | | |
|---|---|---|---|---|---|---|---|---|---|---|
| Patel (2003) | CIS-R, BDQ, GHQ | CIS-R (Psychiatric morbidity) | Response reported | No | No | No | No | No | No | No |
| Fruehwald (2003) | HAMD 17, BDI, CGI, SSS, MMSE, BI | HDS score | Response reported | 18 month follow-up | Limited details (no frequency or statistical analysis) | Yes | No | No | No | Offered continued treatment "for relapse prevention" |
| Rickels (2003) | HAMA, HADS, MADRS, CGI, Sheehan Disability Scale | HAMA score | Yes | No | Yes | Yes | Yes | No | No | No |

| | | | | | | | | | | |
|---|---|---|---|---|---|---|---|---|---|---|
| Mulholland (2003) | BDI, HAMD unspecified, BPRS, SANS, CGI | BDI | Response reported | No | Yes | Yes | "Decreased libido" is mentioned | No | No | No |
| Fisch (2003) | FACT-G , BZSDS, TQSS, Functional Assessment of Chronic Illness Therapy - Spiritual | FACT-G (quality of life) | Response reported | No | Yes | Yes | No | No | No | Option to continue medication for 9 months |
| Paile-Hyvärinen (2003) | RAND-36, HAMA, MADRS, BDI | RAND-36 (quality of life) score | No | No | AE frequency without statistical analysis | Yes | No | No | No | No |
| Leentjens (2003) | MADRS | MADRS Score | Response reported | No | No | Yes | No | No | No | No |

| | | | | | | | | | | |
|---|---|---|---|---|---|---|---|---|---|---|
| Lepola (2003) | MADRS, CGI | MADRS score change | Yes | No | Yes | Yes | Yes | No | No | No |
| Schneider (2003) | HAMD 17, CGI-S, CGI-I, MMSE, Quality of Life Enjoyment and Satisfaction Questionnaire, SF-36, PGI | Hamilton Depression Rating Scale, CGI-S, CGI-I | Response reported | No | Yes | Yes | No | No | No | No |
| Lyketsos (2003) | CSDD, HAMD 17, MMSE, Psychogeriatric Depression Rating Scale-activities of daily living subscale, NPI | Cornell Scale for Depression in Dementia, HDRS | Response reported | No | Yes | Yes | No | No | No | No |

| | | | | | | | | | | |
|---|---|---|---|---|---|---|---|---|---|---|
| Lenox-Smith (2003) | HAMA, HADS, MADRS, CGI, SF-36 | HAMA score | Yes | No | AEs with frequencies, but no statistical analysis | Yes | No | 1 week taper | No | No |
| Moak (2003) | HAMD 21, BDI, OCDS | HAMD; BDI; Drinking outcomes | No | No, 16 & 26 week post-treatment assessment reported separately | AEs generally listed without frequency or statistics | Yes | Decreased libido mentioned as reason for discontinuation once | 1 week taper | No | No |
| Gual (2003) | MADRS, HAMD 17, SF-36 | MADRS Response rate (≥50% reduction in score), relapse in alcohol consumption | Response reported | No | AE frequency reported, but no statistical analysis | Yes | No | No | No | No |
| Rapaport (2003) | HAMD-17, CGI-S, CGI-I | HAM-D total score | Yes | No | AE frequency without statistical analysis | Yes | Abnormal ejaculation >10% of patients in treatment arm | Optional 10-day taper | No | No |
| Swenson (2003) | HAMD 17, CGI-I, BDI, Q-LES-Q, SF-36 | Q-LES-Q | Response reported | No | No | No | No | 2 week taper | No | No, referral for further treatment |

| | | | | | | | | | | |
|---|---|---|---|---|---|---|---|---|---|---|
| Rynn (2008) | HAMA, CGI-S, HADS, CAS, RDS, CGI-I, PGI-I, SDS | HAMA score | Yes | No | Yes | Yes | Yes, decreased libido reported | 2 week taper | DEAEs reported but monitoring window not specified, 2 week taper, referral for continued treatment | No |
| Bose/Korotzer (2008) | HAMA, HAMD 17, HAD, CGI-S, CGI-I | HAMA score | Yes | No | Yes | Yes | Yes | 2 week taper | No | No |
| Navari (2008) | BZSDS, TQSS | Depressive symptoms, chemotherapy completion, quality of life | No | No | No AEs | Yes | No | No | No | No |
| Kraus (2008) | HADS | HADS score | No | 4w follow-up | No AEs | Yes | No | No | No | No |
| Bose/Li (2008) | MADRS, HAMD 17, HAM-D 24, CGI-S, CGI-I | MADRS score | Yes | No | Yes | Yes | No | No | No | No |

| | | | | | | | | | | |
|---|---|---|---|---|---|---|---|---|---|---|
| Ehde (2008) | HAMD 17, CES-D, SCL-20, HAMA | HAMD response | Yes | No | Frequency of AEs reported without statistical analysis | Yes | Yes | No | No | Offered continued medication with PCP |
| Raskin (2008) | HAMD 17, Geriatric Depression Scale, CGI-S | Protocol-specified composite cognitive score | No | No | Yes | Yes | Yes (libido) | 1 week taper | DEAEs reported (duloxetine 17.3%, placebo 11.3%), but unclear follow-up interval | No |
| Yonkers (2008) | HAMD 17; IDS-SR; CGI-I, CGI-S, SCID | HAMD-17 score and remission; IDS-SR; CGI Scores and response (CGI-I) | Yes | No | Yes | Yes | Yes (“sexual dysfunction” | Yes (1-week) | No | No |
| Levin (2013) | HAMD 17, CGI | HAMD-17 score, MJ use | Yes | No | Yes | Yes | Yes (“loss of libido”) | No | No | No |
| Banerjee (2013) | CSDD, HAMD (for diagnosis) | CSDD score | No | No | Yes | Yes | Yes | No | No | Participants given 4 additional weeks of medication at completion |

| | | | | | | | | | | |
|---|---|---|---|---|---|---|---|---|---|---|
| Ravindran (2013) | HAMD 17, CGI-S, CGI-S, BDI, Q-LES-Q | Change in HAMD-17 | Yes | No | Yes | Yes | Yes (problems related to arousal, ejaculation, libido, orgasm) | No | No | No |
| Brown (2018) | HAMD 17; IDS-SR, CGI-S, CGI-I, Quality of Life Enjoyment and Satisfaction Questionnaire | HAMD17; IDS-SR | No | No | Yes | Yes | No | No | No | No |
| Valle-Cabrera (2018) | HAMD 17, CGI-I, CGI-S | HAMD17 response | Yes | No | Yes | Yes | No | No | No | Participants offered 6 months open-label treatment at completion |

Supplementary Table 2 Abbreviations: AEs: Adverse Events; AMDP: Association for Methodology and Documentation in Psychiatry; ASI: Addiction Severity Index; BDI: Beck Depression Inventory; BDQ: Brief Disability Questionnaire; BI: Barthel Index; BPRS: Brief Psychiatric Rating Scale; BZSDS: Brief Zung Self-Rating Depression Scale; CAS: Covi Anxiety Scale; CES-D: Center for Epidemiologic Studies Depression Scale; CGI-I: Clinical Global Impression-Improvement; CGI-S: Clinical Global Impression-Severity; CIS-R: Clinical Interview Schedule-Revised; CSDD: Cornell Scale for Depression in Dementia; DEAE: Discontinuation-Emergent Adverse Events; FACT-G: Functional Assessment of Cancer Therapy-General; GHQ: General Health Questionnaire; GP: General Practitioner; HADS: Hospital Anxiety and Depression Scale; HAMA: Hamilton Anxiety Rating Scale; HAMD/HDRS: Hamilton Depression Rating Scale; IDS-SR: Inventory of Depressive Symptomatology-Self Report; MADRS: Montgomery-Åsberg Depression Rating Scale; MMSE: Mini-Mental State Examination; NPI: Neuropsychiatric Inventory; OCDS: Obsessive Compulsive Drinking Scale; PCP: Primary Care Physician; PGI/PGE: Patient Global Impression/Patient Global Experience; Q-LES-Q: Quality of Life

Enjoyment and Satisfaction Questionnaire; RADS: Raskin Depression Scale; RAND-36: RAND 36-Item Health Survey; SANS: Schedule for the Assessment of Negative Symptoms; SCID: Structured Clinical Interview for DSM; SCL-20: Hopkins Symptom Checklist (20-item version); SDS: Sheehan Disability Scale; SF-36: 36-Item Short Form Health Survey; SSS: Scandinavian Stroke Scale; TQSS: Two-Question Screening Survey.
